# Assessing Swedish Genetic Counselling Outcome Measures for Autism and General Use: Rasch Findings Highlight the Need for Improved Measures

**DOI:** 10.64898/2026.04.13.26350766

**Authors:** Mattias Nordstrand, Samuelle Fajutrao Falk, Magnus Johansson, Rebecka Pestoff, Kristiina Tammimies

**Author notes:** Corresponding authors: Samuelle Fajutrao Falk, and Kristiina Tammimies, Biomedicum A04/Tammimies, Karolinska Institutet, 171 77 Stockholm, Sweden. These authors contributed equally (Mattias Nordstrand, Samuelle Fajutrao Falk).

## Abstract

Genetic counselling outcome measures are increasingly adapted for diverse clinical contexts. While the Genetic Counselling Outcome Scale (GCOS-24) is available in Swedish, no autism-specific version has been developed. Therefore, we adapted the Swedish GCOS-24 using the English version of the modified GCOS-24 (mGCSOS-24) to create a Swedish autism-specific mGCOS-24. Thereafter, we evaluated both the Swedish autism mGCOS-24 and the Swedish general GCOS-24 using Rasch analysis to assess their psychometric properties. Both instruments exhibited structural challenges, including multidimensionality, disordered thresholds, local item dependence, and invariance issues. For the Swedish autism mGCOS-24, we were able to identify subscales with acceptable measurement properties. However, applying the same structure to the Swedish general GCOS-24 did not resolve its broader limitations. This study introduces the first Swedish autism-specific mGCOS-24 and represents the first Rasch-based evaluation of any GCOS-24 or mGCOS-24 in Swedish. Our findings highlight important opportunities for measure refinement but also indicate that new or more substantially adapted tools may be needed to capture outcomes of genetic counselling in autistic populations.

## Introduction

Autism is a neurodevelopmental condition with a strong genetic component. Twin and family studies estimate heritability between 64% and 91%, and children of autistic parents have an increased likelihood of being autistic compared with children of non-autistic parents (1–3). Because of this strong genetic background, families of autistic children often seek information about genetic causes, inheritance, and implications for their child’s future health (4). Genetic counselling, whether provided with or without a genetic test result, can support families in understanding genetic concepts, interpreting information, and adapting to the implications of genetic vulnerability (5). Although genetic testing for those with autism diagnosis has been recommended in several clinical guidelines (6,7), access to testing remains limited in many settings, including in Sweden, where awareness of and access to clinical genetic testing for autism are still relatively low (Hellquist & Tammimies, 2022). In this context, genetic counselling may play an important role regardless of whether genetic testing is performed or yields a definitive result.

Genetic counselling aims to support patients in understanding, interpreting, and adapting to genetic information, thereby facilitating informed decision-making and psychological adjustment (9). In the context of autism, genetic counselling has been proposed as a way to help individuals and families better understand the genetic aspects of the condition and navigate associated uncertainties (10). However, genetic counselling for autism remains relatively limited globally, and its impact has not been extensively evaluated. Reliable outcome measures are therefore needed to assess whether genetic counselling interventions provide meaningful benefits for individuals and families affected by autism.

Patient-reported outcome measures (PROMs) play a central role in evaluating healthcare interventions by capturing patients’ perspectives on their experiences and outcomes (11,12). In the context of genetic counselling, outcomes of interest include knowledge, satisfaction, psychological well-being, perceived risk, decisional conflict, and empowerment (13). Among these, empowerment has been identified as a particularly relevant outcome, as it reflects individuals’ perceived control, understanding, emotional regulation, and capacity to make informed decisions in the context of genetic vulnerabilities (14).

The Genetic Counselling Outcome Scale (GCOS-24) was developed to measure patient empowerment, based on research with patients and genetics service providers. It assesses five key aspects of empowerment: behavioral control, decisional control, cognitive control, emotional regulation, and hope (13). The GCOS-24 has been translated, culturally adapted, and validated across several cultural contexts, including in Swedish populations (15–24).

Despite its broad use, the measurement properties of GCOS-24 have not been fully studied. Most validation studies on GCOS-24, including the Swedish GCOS-24 (20), have relied on classical test theory (CTT) and factor analysis, which primarily evaluate instruments based on overall scale properties such as reliability and item–total correlations. While these approaches can provide useful information about internal consistency and factor structures, they offer limited insight into the functioning of individual items, including their consistency across different subpopulations. Additionally, the use of rule-of-thumb critical values to interpret model fit metrics in factor analysis is problematic and may frequently lead to incorrect conclusions (25–27). In contrast, Rasch measurement theory (RMT) provides a model-based framework that assesses whether observed responses fit the expectations of a measurement model. This approach enables detailed examination of item functioning, local independence of items, response category structure, dimensionality, and measurement invariance, thereby supporting the development of valid and interpretable measurement scales (28,29). To date, only a limited number of studies have applied RMT to evaluate GCOS-24. These studies have reported mixed findings and identified potential challenges related to dimensionality, item functioning, and response category structure, which may limit the interpretability and comparability of GCOS-24 scores across populations and contexts (15,17,24). It is also important to note that previous RMT studies have used software that relies on unconditional item fit, which has been found to be unreliable with sample sizes above 200 (30), and that, similarly to CTT, rule-of-thumb critical values are likely to lead to incorrect interpretations of results, for instance regarding for eigenvalues from principal component analyses of residuals (31) and residual correlations between items (32).

Another consideration is that GCOS-24 was designed as a general instrument intended to be applicable across different genetic conditions and clinical settings. For general instruments, it is assumed that the construct of interest can be measured in a similar manner across conditions and populations. However, this assumption may not always hold. For example, the interpretation of empowerment-related items may differ depending on the clinical context, the nature of the condition, and the experiences of the individuals completing the measurement. Therefore, it is important to examine how GCOS-24 performs in specific contexts such as autism.

To address this issue, one study adapted the GCOS-24 to create a modified version (mGCOS-24) specifically for parents of autistic children in English and French (33). The modified scale demonstrated promising initial evidence of construct validity. However, no study has yet conducted a comprehensive Rasch-based evaluation of a Swedish autism-adapted version of GCOS-24, nor examined whether the measurement properties of the Swedish GCOS-24 extend to autism-related contexts beyond the initial validation (20).

This present study aims to evaluate the measurement properties of the Swedish autism-adapted version of the scale, as well as the Swedish general GCOS-24 for comparison, using Rasch measurement theory. First, we translate and adapt the Swedish GCOS-24 to develop a Swedish autism mGCOS-24. We then applied Rasch measurement theory, complemented by classical reliability indices, to examine dimensionality, item functioning, response category functioning, targeting, and reliability. Based on these analyses, we explored potential improvements to the scale, including the formation of subscales and the collapsing of response categories where appropriate. Finally, we evaluated whether similar structural limitations are present in both the Swedish mGCOS-24 and the Swedish general GCOS-24, and whether derived subscales can be applied across populations while maintaining acceptable psychometric validity.

## Methods

### Adaptation of the Swedish GCOS-24 for autism and other neurodevelopmental conditions

We adapted the Swedish translation of the GCOS-24 measure, previously developed by Pestoff and colleagues, to better suit autistic and other neurodevelopmental populations (20). The adapted scale is available in **Table S 1**. We adapted the scale similarly to how the English version had earlier been adapted for autism (33).

References to “medical condition” were replaced with “neurodevelopmental condition” or removed to avoid medicalizing language. Items were revised to include family perspectives, reflecting that participants may be family members attending genetic services. First-person phrasing was added where applicable, allowing individuals with the condition to complete the measure directly. An item on seeking available support was revised to include psychological support, reflecting services commonly offered for patients in this population.

References to control programs and prenatal testing were removed, as these are not relevant in a Swedish context for this population group. Many other items were adapted consistently with prior modifications.

### Recruitment of the autistic participants

Participants answering the Swedish autism mGCOS-24 were recruited between March and August 2024 from two primary sources: (1) a previously established cohort of autistic individuals who had taken part in the “KONTAKT” social skills randomized controlled trial (34), and (2) additional autistic individuals recruited through patient organizations, social media channels, and outpatient psychiatric clinics. A total of 55 participants aged 15-30 years old were recruited via the first source and registered their responses through the technical platform and database (BASS4) provided by Karolinska Core Facilities. This subset of participants had been recruited as part of a clinical trial evaluating a genetic information intervention (ClinicalTrials.gov ID NCT06740760, 05/08/2025). As per the second source, a total of 118 autistic adults (≥18 years) had their responses collected in the Qualtrics online tool (35). Participants from both sources were combined into a single study sample. In total, this included 171 responses where at least two questions were answered, and 155 were complete responses.

### GCOS-24 clinical genetics sample

When examining the Swedish general GCOS-24, we reused the data that was collected for the original study on the translation, cross-cultural adaptation, and preliminary validation of the Swedish version of the GCOS-24 (20). In total, 344 participants responded to all questions with 383 responding to at least two of the questions prior to genetic counselling and the subsequent assessment.

In brief, for this sample, participants were recruited from those scheduled for genetic counselling at clinical genetics departments at either Linköping University Hospital or the Karolinska University Hospital, in Sweden, and the participants were asked to complete the Swedish GCOS-24 before and after their genetic counselling session. The sample included individuals with a wide range of medical conditions with majority within the cancer care, reflecting the heterogeneity of routine clinical genetics practice. In our study, we specifically used the data subset that was collected prior to the genetic counselling session through digital consultation (video/telephone). Detailed information on recruitment procedures, inclusion criteria, clinical settings, and data collection methods is reported in (20).

### Rasch Analysis and Preprocessing

As for pre-processing the data, participants included were those with two or more answers to the items included in the scale/subscale. Participants with invalid responses outside the Likert scale (1 to 7) were not included. The Likert scales of negatively phrased questions (item 4, 11, 21, and 22) were inverted prior to the analysis (**Table S1**). The response numbers (scale options) were further shifted to become 0-indexed. Additional details on the preprocessing are described in the Supplementary Method section.

Our psychometric evaluation followed the minimal checklist for Rasch-based scale assessment (36) including unidimensionality, local independence of items, monotonically ordered response categories, invariance, targeting, and reliability. Unidimensionality and local dependencies were examined using conditional information weighted mean-square item fit statistics, “infit MSQ” (30), residual correlations between item pairs (32), and eigenvalues from principal component analysis of Rasch residuals (31). These metrics were assessed against simulation-based critical values (37), which are determined by using the data being analyzed to generate a large amount of similar, simulated datasets that fit the measurement model.

Targeting and response category functioning were assessed through visual inspection of person and item location distributions and category threshold ordering. Measurement invariance was examined using Andersen’s likelihood ratio (LR) test (38) with respect to age and sex. Reliability was evaluated using Cronbach’s alpha, the Person Separation Index (PSI), and a proposed new metric, Relative Measurement Uncertainty (WLE-RMU) (39).

Data analysis was conducted in R, version 4.5.1, using easyRasch, version 0.4.2, which acts as a wrapper for related packages such as eRm, version 1.0.10 (40), iarm (41), and mirt (42), version 1.45.1. We performed the analyses twice, with and without imputation using the R package mice (43).

### Subscales and Categories

We followed the recommended steps (36) for when a scale does not fulfill basic psychometric properties. Whenever there was multidimensionality, we divided the scale into unidimensional subscales. If the scale had disordered thresholds for any item following Rasch, we collapsed the adjacent response categories as needed to create fewer, more distinct response categories and obtain ordered thresholds.

We designed subscales as needed by grouping items thematically while avoiding the inclusion of highly similar items within the same subscale. The subscale compositions were evaluated iteratively using Rasch analysis to examine their psychometric validity until we found a working set of subscales. Heuristically, we then combined each pair of subscales. If resulting combinations remained multidimensional, we concluded that each subscale measured a distinct latent construct and ceased attempts to form larger subscales.

First, we followed this process for the Swedish autism mGCOS-24 data. We then evaluated the same subscales for the Swedish general GCOS-24 data to evaluate whether the subscales were generalizable between populations and minor scale variations.

## Results

### Participants

Following the pre-processing, the Swedish autism mGCOS-24 group consisted of 171 participants, and the Swedish general GCOS-24 consisted of 383 participants (**Table 1**). In both cohorts, most participants were female (77% and 74%, respectively). The autism mGCOS-24 group included only individuals diagnosed with autism, whereas the general GCOS cohort represented a heterogeneous group of conditions (including autism), with nearly half of the participants having cancer. Both cohorts spanned a wide age range with a median age of 28 years in the autism mGCOS-24 group and 47 years in the general GCOS-24 group (**Table 1**).

**Table 1.** Demographic and clinical characteristics of participants completing the Swedish autism mGCOS-24 and the Swedish general GCOS-24.

| **Scale** | **Participants** | **Age** | **Sex** | **Conditions** |
| --- | --- | --- | --- | --- |
| **Swedish Autism mGCOS-24** | 171 (of which 155 answered all items) | Mean: 30.94 years<br>Median: 28 years<br>Min: 16 years<br>Max: 62 years<br>Std: 10.0 | M: 39 (23%)<br>F: 131 (77%) | 171 autism (100%) |
| **Swedish General GCOS-24** | 383 (of which 344 answered all items) | Mean: 46.93 years<br>Median 47 years<br>Min: 2 years<br>Max: 89 years<br>Std: 16.76 | M: 100 (26%)<br>F: 281 (74%) | 189 cancer (49%),<br>194 other (51%) |

### Psychometrics – Entire Scales

The original Swedish autism mGCOS-24 demonstrated acceptable internal consistency and separation reliability (Cronbach’s α = 0.84, PSI = 0.852), but also showed substantial measurement deficits, including multidimensionality (**Figure 1 A-B, Table 2**). Principal component analysis of the residuals yielded a first eigenvalue of 5.08, exceeding both commonly used rule-of-thumb thresholds for unidimensionality (44) as well as the simulated threshold of 2.06. Rasch model fit statistics further indicated misfit across most items (**Table S 2**), with infit MSQ values falling outside the simulated acceptable interval, reflecting both overfit and underfit. There were also many residual correlations above the simulated cutoff (**Figure S 1**). Together, these findings indicate that the Swedish autism mGCOS-24 is not a unidimensional measurement scale.

**Table 2.** Summary of Rasch model diagnostics and reliability indices for the Swedish autism mGCOS-24 and Swedish general GCOS-24, including entire scales and derived subscales.

| Scale | First Eigenvalue for Residuals | PSI | WLE-RMU | Cronbach's alpha | Age LR-test (p) | Sex LR-test (p) |
| --- | --- | --- | --- | --- | --- | --- |
| **Swedish Autism mGCOS-24** |  |  |  |  |  |  |
| Entire Scale | 5.08 (Sim. Threshold 2.06) | 0.852 | 0.848 (95% HDCI [0.814, 0.882]) | 0.84 (95% CI [0.81,0.88]) | 0.026 | 0.133 |
| Subscale 1 | 1.56 (Sim. Threshold 1.60) | 0.552 | 0.562 (95% HDCI [0.467, 0.651]) | 0.55 (95% CI [0.44,0.66]) | 0.092 | 0.098 |
| Subscale 2 | 1.51 (Sim. Threshold 1.71) | 0.757 | 0.795 (95% HDCI [0.749, 0.838]) | 0.79 (95% CI [0.74,0.84]) | 0.787 | 0.619 |
| Subscale 3 | 1.60 (Sim. Threshold 1.73) | 0.683 | 0.782 (95% HDCI [0.734, 0.829]) | 0.77 (95% CI [0.71,0.83]) | 0.741 | 0.445 |
| Subscale 4 | 1.61 (Sim. Threshold 1.70) | 0.758 | 0.768 (95% HDCI [0.718, 0.817]) | 0.76 (95% CI [0.7,0.81]) | 0.108 | 0.429 |
| **Swedish General GCOS-24** |  |  |  |  |  |  |
| Entire Scale | 4.80 (Sim. Threshold 1.65) | 0.867 | 0.863 (95% HDCI [0.843, 0.883]) | 0.86(95% CI [0.84,0.88]) | < 0.001 | < 0.001 |
| Subscale 1 | 1.70 (Sim. Threshold 1.47) | 0.612 | 0.646 (95% HDCI [0.596, 0.695]) | 0.63 (95% CI [0.58,0.69]) | 0.019 | 0.010 |
| Subscale 2 | 1.79 (Sim. Threshold 1.49) | 0.730 | 0.757 (95% HDCI [0.722, 0.791]) | 0.75 (95% CI [0.70,0.79]) | 0.044 | 0.720 |
| Subscale 3 | 1.64 (Sim. Threshold 1.64) | 0.553 | 0.724 (95% HDCI [0.686, 0.763]) | 0.72 (95% CI [0.67,0.77]) | 0.192 | 0.110 |
| Subscale 4 | 1.62 (Sim. Threshold 1.47) | 0.780 | 0.783 (95% HDCI [0.752, 0.814]) | 0.77 (95% CI [0.74,0.81]) | 0.155 | 0.005 |

**Figure 1.**
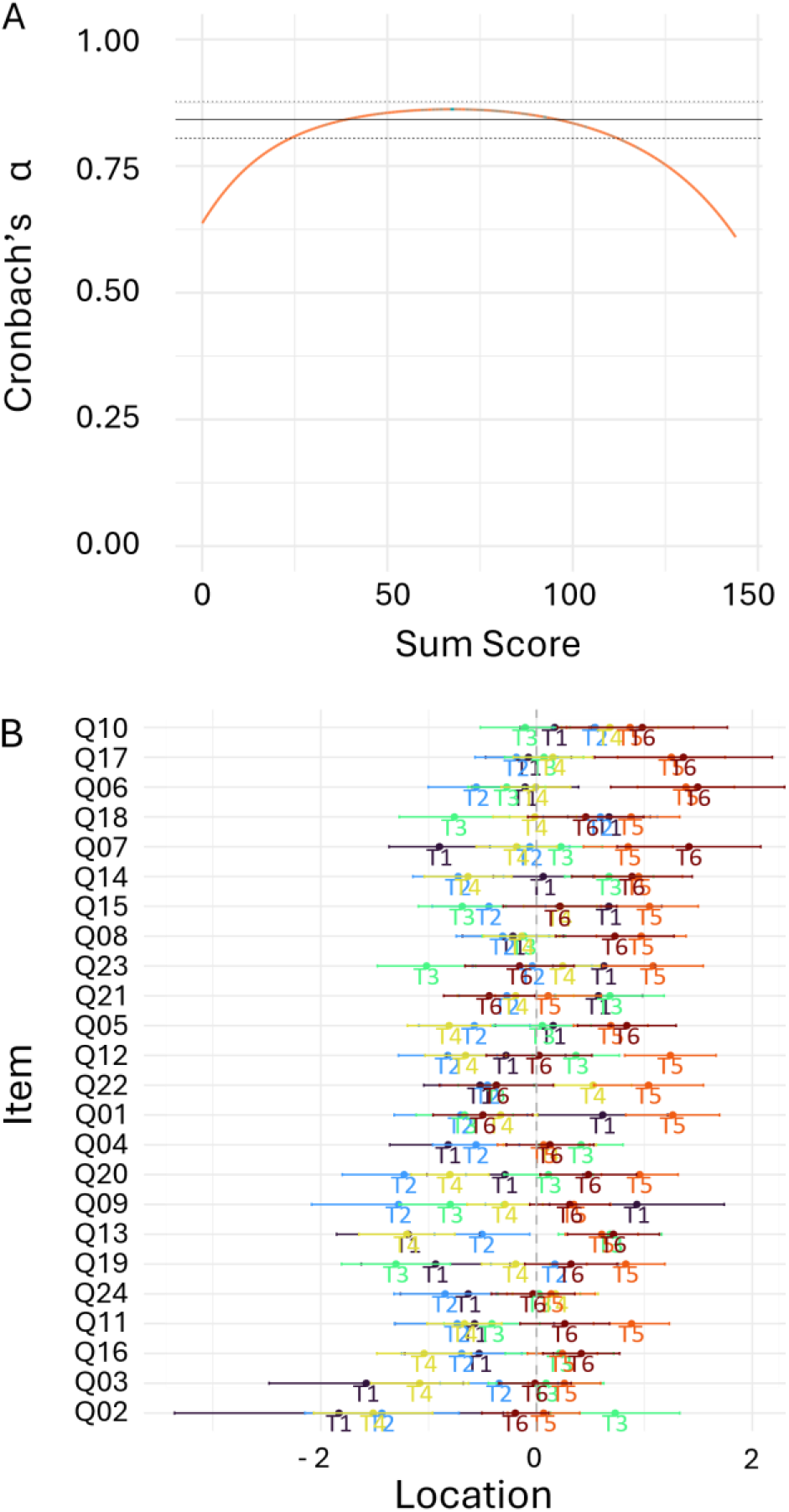
Rasch diagnostic outputs for the full Swedish autism mGCOS-24 scale. (A) Internal consistency plot showing the point estimate of Cronbach’s alpha, the confidence interval and the reliability curve of alpha across sum scores. (B) Category threshold locations illustrating disordered thresholds across all response categories, indicating that adjacent response options did not operate in the intended ordinal sequence. Ti denotes the estimated location of the i-th category threshold along the latent trait continuum.

Moreover, there were disordered response thresholds after fitting the Rasch model (**Figure 1 C**). This pattern indicates that the ordering of response categories did not function as expected, with some adjacent categories failing to show distinct transitions along the latent trait continuum. The presence of threshold disordering was consistent across all items rather than being restricted to a single item. Furthermore, the scale was not invariant with respect to age (**Table 2)** with significant differential item functioning detected between age groups.

The psychometric pattern observed for the Swedish autism-adapted mGCOS-24 closely mirrored that of the Swedish general GCOS-24 (**Table 2, Table S 3, Figure S 2**). Similarly, the general GCOS-24 demonstrated multidimensionality, disordered response thresholds, local dependency, and lack of age invariance. In addition, the general GCOS-24 showed non-invariance with respect to sex. Taken together, these findings indicate a highly similar measurement structure and set of limitations across the two Swedish versions.

### Psychometrics – Subscales

After iteratively restructuring the Swedish autism mGCOS-24 into subscales and collapsing adjacent response categories to achieve ordered response thresholds, the final structure consisted of four subscales that we named “Knowledge”, “Hope”, “Coping” and “Cognitive control” **(Figure 2 A)**. Items 4, 19 and 23 were excluded from the subscales due to persistent item misfit and residual correlations. The subscales were named and organized to align with those in a previous study identifying similar themes and subdomains, although they were based on partially different items (17). Collapsing scale response categories 2 to 4 (“Disagree”, “Slightly disagree”, and “Neither agree nor disagree”) into a single category and 5 to 6 (“Slightly agree” and “Agree”) into another, with additional merging of the two now lowest categories (“Strongly disagree” and “Disagree”/“Slightly disagree”/“Neither agree nor disagree”) for items 2 and 3 for the Swedish autism mGCOS-24 due to skewed response distribution (see Supplementary Method), resulted in subscales demonstrating ordered thresholds, unidimensionality, and adequate targeting (**Figure 2 B-E, Table 2**).

**Figure 2.**
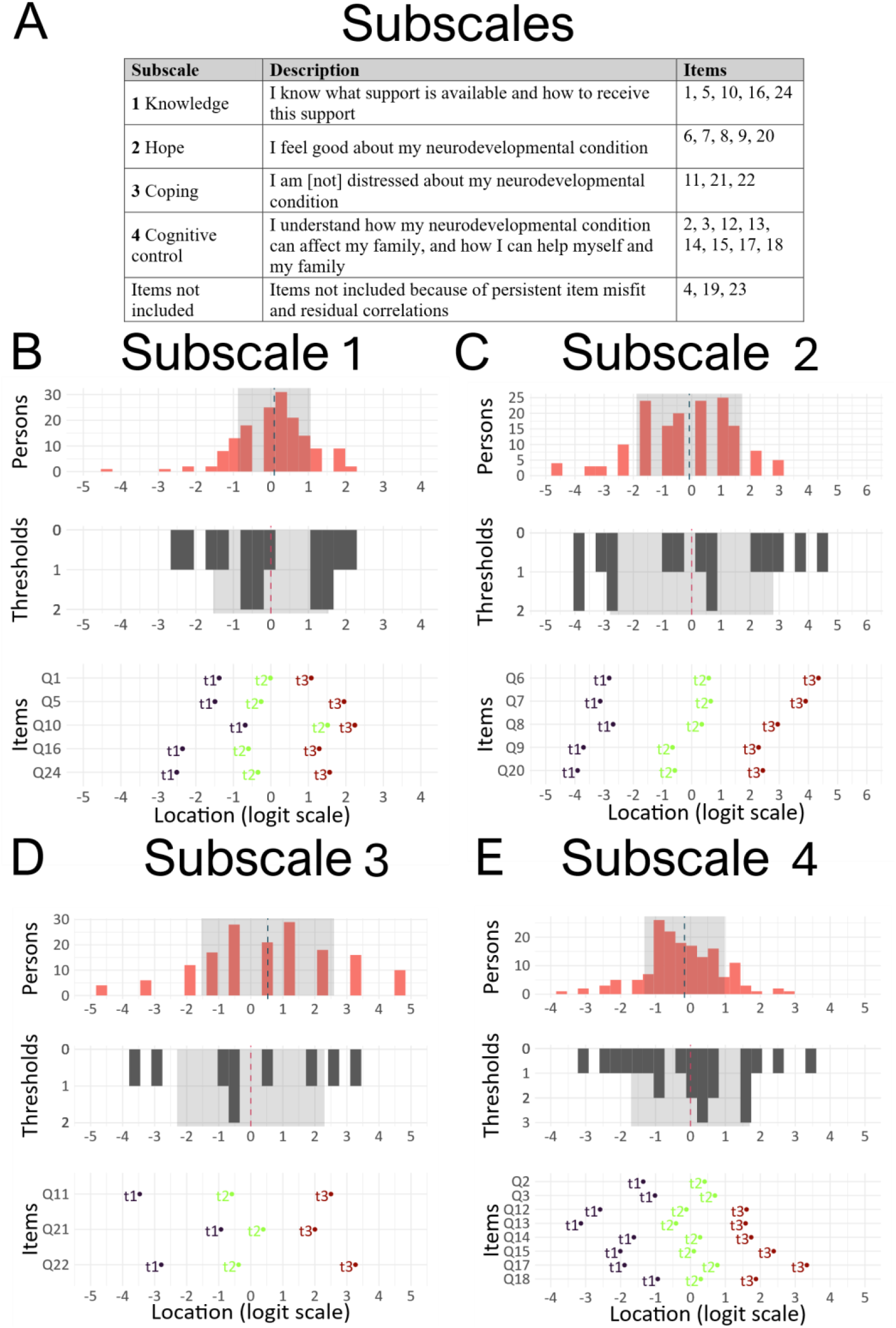
Targeting and category functioning for the four finalized Rasch-validated subscales of the Swedish autism mGCOS-24. (A) Subscale composition, listing assigned items and subscale structure. (B–E) Person, threshold and item locations for each subscale, showing adequate targeting between item difficulty and participant trait levels, as well as ordered category thresholds after category collapsing. Sample size across panels ranges from n = 156–166, depending on item-level completeness.

For all subscales, the first eigenvalue was smaller than the simulation-based eigenvalue thresholds (**Table 2**). Item misfit per item was minimal (maximum infit MSQ deviation = 0.044 from the simulated threshold), and no residual correlations exceeded the simulated cutoff. These findings were consistent both with and without imputation of missing data prior to the Rasch analyses. Combining the subscales (**Table S 4**), in terms of all possible pairs of item sets, leads to multidimensionality. Correlations of person locations between subscales ranged from negligible (r = -0.119, not significant) to moderate (r = 0.688, p < 0.001) (**Figure S 3**). This indicates that the subscales do not share a common latent construct but instead have distinct constructs. Furthermore, invariance was assessed using Andersen’s likelihood ratio test (Table 2). No statistically significant differences were observed for sex or age for any of the subscales of the Swedish autism mGCOS-24.

Across the subscales, item location distributions showed no pronounced ceiling effects or gaps, and person and item locations were well aligned, indicating good targeting (**Figure 2 B-E**). Reliability estimates ranged from low for subscale 1(“Knowledge”) to medium for subscales 2-4 (“Hope”, “Coping” and “Cognitive control”) (**Table 2**).

We then investigated whether the subscales found for the Swedish autism mGCOS-24 could generalize to the Swedish general GCOS-24 and its corresponding participant population. The validity of the subscales from the Swedish autism mGCOS-24 did not generalize very well to the Swedish general GCOS-24, except possibly for subscale 3 (“Coping”), which remained essentially unidimensional. For the other subscales (“Knowledge”, “Hope” and “Cognitive control”), there were clear indications of invariance issues and indications of multidimensionality for the Swedish general GCOS (**Table 2**). Moreover, the Genomic Outcome Scale, a subscale of GCOS-24 previously proposed based on Rasch analysis (17), is also multidimensional in both the Swedish autism mGCOS-24 and the Swedish general GCOS-24 using the current data (**Table S 4)**. Thus, there are no indications that subscales generalize well between study populations or minor scale modifications.

## Discussion

As genetic counselling, with or without genetic testing, is increasingly used in both clinical practice and research, reliable PROMs are needed to assess its effects. In this study, we evaluated the psychometric properties of the Swedish autism mGCOS-24 using RMT. While Cronbach’s alpha suggested acceptable performance of the total scale, Rasch-based analyses revealed violations of unidimensionality, indicating that both the Swedish autism mGCOS-24 and the Swedish general GCOS-24 capture multiple distinct latent constructs rather than a single overarching dimension of empowerment.

Although the GCOS-24 and autism mGCOS-24 were originally designed as single scales, the observed multidimensionality is consistent with the multiple conceptual domains originally proposed for the GCOS-24 (13). When the Swedish autism mGCOS-24 was divided into themed subscales, Rasch model assumptions were met, response categories were ordered, unidimensionality was supported, local independence was maintained, and invariance across age and sex indicated stable item functioning. Targeting was also adequate across subscales.

While these subscales performed well for the Swedish autism mGCOS-24, they did not generalize to the Swedish general GCOS-24. Although a universal outcome measure applicable across conditions would be desirable, achieving this while maintaining invariance may be challenging, as items may be interpreted differently depending on the underlying condition. This can introduce construct-irrelevant variation, threatening invariance and reducing comparability between groups. Notably, without subscales, the Swedish autism mGCOS-24 was not invariant to age despite including only individuals with autism, suggesting age-related influences on responses beyond the latent construct. This undermines the interpretability of total scores, as observed differences may reflect bias rather than true variation. Future development of genetic counselling outcome measures should therefore explicitly address invariance, particularly if instruments are intended for use across conditions or demographic groups.

We also observed problems related to disordered Rasch thresholds, meaning that transitions between response options did not follow the expected progression along the latent variable. All three previous Rasch analyses of the GCOS-24 also found similar issues. One likely explanation is that the labels may be similar enough to make adjacent categories hard to distinguish from each other in a meaningful way. Another issue is that both the Swedish autism GCOS-24 and the Swedish general GCOS-24 instruct respondents to select the middle option (“neither agree nor disagree”) when an item feels irrelevant. This is incompatible with the ordinal nature of the response scale. A separate response option for “irrelevant”, or allowing respondents to skip non-applicable items, would likely improve measurement clarity. In the present study, we collapsed adjacent categories to obtain ordered thresholds in the subscales, but this type of post hoc adjustment might not be necessary if response formats and instructions were designed with category functioning in mind from the beginning.

These psychometric shortcomings have important implications for the use of GCOS-24-based measures as outcome instruments in genetic counselling research. The observed multidimensionality indicates that empowerment is a broad and multifaceted construct encompassing several dimensions (45). Although summarizing responses into a single score may be appealing, this reduces conceptual clarity and limits the ability to detect how counselling impacts specific aspects of empowerment. To more accurately capture these effects and enhance clinical interpretability, results should therefore be presented at the subdomain level, which is likely to provide more precise and clinically meaningful insights. If reporting an overall score is nevertheless desired for practical reasons, results should be presented alongside subdomain scores, and a more transparent approach would be to standardize subscale scores and average them across domains to obtain an overall empowerment index. However, such an index should be interpreted with caution, as it would not represent a unidimensional Rasch-based measure and would require further methodological work to establish appropriate standardization procedures.

As for translations and condition-adaptations of the GCOS-24, Cronbach’s alpha has been one important metric in many GCOS-modification studies (16,18–20,22,33). Our study indicates that traditional methods, such as Cronbach’s alpha, are insufficient to properly evaluate psychometric properties of a scale and should be complemented by modern, comprehensive psychometric analyses, such as those provided by RMT.

Our results largely align with previous Rasch-based evaluations of the GCOS-24, which have reported inconsistent psychometric performance and highlighted challenges related to dimensionality and item functioning (15,17,24). The present study extends this work by demonstrating that, in autistic individuals, measuring specific aspects of empowerment yields greater transparency and usability than attempting to capture all components of empowerment into a single summary score. This is particularly relevant given the heterogeneity of autistic experiences and the likelihood that genetic counselling impacts cognitive, emotional, and motivational domains in different ways. A key difference compared to earlier studies was that of the Genomic Outcome Scale suggested by (17), which we found to be multidimensional. Apart from the possibility of language and study sample differences, our use of simulation-based critical values and conditional item fit statistics are likely to produce different and more accurate results.

The study is limited primarily by sample size, particularly for the Swedish autism mGCOS-24. Sample size influences both Rasch model fit and the interpretation of invariance tests (37). For example, the Andersen LR test requires subgrouping, which reduces power substantially in smaller samples. Replication with larger, independent datasets will therefore be essential to confirm the dimensional structure identified here and to refine the proposed subscales of the Swedish autism mGCOS-24.

In addition, recruitment methods may have introduced bias. Online recruitment for the autistic cohort may have favored individuals who are more engaged, more comfortable responding digitally, or more motivated to participate in autism research. Differences in context between participants responding hypothetically versus those entering a genetic counselling trial may also have influenced interpretation of items. Finally, the overrepresentation of female participants may limit the generalizability of the findings, potentially emphasizing perspectives that differ systematically from those of males or nonbinary individuals.

In conclusion, despite acceptable internal consistency, Rasch analysis indicates that neither instrument functions as a unidimensional measure, instead capturing multiple subdomains. Subscales for the Swedish autism mGCOS-24 showed acceptable properties but did not generalize to the Swedish general GCOS-24, underscoring the importance of context-specific measurement and the need to develop more robust outcome measures for genetic counselling in the future.

## Data and code sharing

The anonymized Swedish Autism mGCOS-24 data will be available via the KI Data Repository after necessary access clearance. The general Swedish GCOS-24 data can be requested from Rebecka Pestoff.

The code used to generate the results and figures is available on GitHub https://github.com/Tammimies-Lab/GCOS-24_RaschAnalysis_2026.

## Data Availability

The anonymized Swedish Autism mGCOS-24 data will be available via the KI Data
Repository after necessary access clearance.
The general Swedish GCOS-24 data can be
requested from Rebecka Pestoff
The code used to generate the results and
figures is available on GitHub.

https://github.com/Tammimies-Lab/GCOS-24_RaschAnalysis_2026.

## Acknowledgements

We thank all the individuals who have answered the questionnaires; without them, this study would not be possible. This project received funding from the LifeWatch Niclas Öberg Foundation (K.T) and Karolinska Institutet Committee for Research (K.T).

## Author contributions

**Mattias Nordstrand**: formal analysis; investigation; methodology; resources; software; validation; visualization; writing – original draft; writing – review and editing.

**Samuelle Fajutrao Falk**: conceptualization; data curation; investigation; methodology; project administration; resources; writing – original draft; writing – review and editing.

**Magnus Johansson**: methodology; resources; validation; writing – review and editing.

**Rebecka Pestoff**: data curation; resources; writing – review and editing.

**Kristiina Tammimies**: conceptualization; funding; methodology; project administration; resources; supervision; writing – review and editing.

## Ethical approval

This study was approved by the Swedish Ethical Review Authority (Dnr. 2023-06737-01; 2024-04937-02). A separate ethical approval was obtained for the Swedish GCOS-24 study (Dnr. 2019-01051; 2020-05243).

## Competing interests

The authors have no COIs related to the work presented here. K.T. is an associate editor for npj Genomic Medicine within the Nature Publishing Group.

## Notes

### Clinical Trial

NCT0674060

### Funding Statement

This project received funding from the LifeWatch
Niclas Oberg Foundation (K.T) and
Karolinska Institutet Committee for
Research (K.T).

### Author Declarations

Ethical Review Authority (Dnr. 2023-06737-01; 2024-04937-02). A separate ethical approval was obtained for the Swedish GCOS-24 study (Dnr. 2019-01051; 2020-05243).

