## Supplementary Files for "Assessing Swedish Genetic Counselling Outcome Measures for Autism and General Use: Rasch Findings Highlight the Need for Improved Measures"

**Supplementary file - Nordstrand, Fajutrao Falk et al.**

**Supplementary methods / Tables /Figures**

Entire scales

The Swedish autism mGCOS (as presented in Table S 1) and the Swedish general GCOS both have large issues with item fit and residual correlations. Below we show the misfit per question for the Swedish autism mGCOS (Table S 2) and the Swedish general GCOS (Table S 3), which indicates multidimensionality for both scales. Furthermore, we show the above-cutoff residual correlations for the Swedish autism mGCOS (Figure S 1) and for the Swedish general GCOS (Figure S 2), which indicates many local dependencies across both scales.

Additional subscales

We combined pairs of subscales and found indications of multidimensionality for all subscale combinations (Table S 4). Specifically, principal component analysis of residuals revealed above-threshold eigenvalues for each subscale pair. Furthermore, we analyzed the person location correlations between subscales for the Swedish autism mGCOS. Some subscales had significant correlations of person locations while others did not. Pearson's r correlation ranged from none (not significant) to moderate-high with the highest correlation being between subscale 1 and subscale 4 (Pearson's r = 0.688).

In addition, the first eigenvalue for the GOS subscale clearly exceeded its simulated threshold (Table S 4). We also observed misfits for several items in GOS (Table S 5), in particular item four. This indicates multidimensionality of GOS under the current data.

Methods details

We did the required preprocessing of inappropriate response patterns when this was necessary. When creating the subscales, if there were fewer than four responses to the lowest response category (after the collapsing), we did an additional merger of the two (new) leftmost response categories to turn the scale into a three-point Likert scale – for those items in that set only. In practice, this only affects item 2 and 3 within subscale 4 for the Swedish autism mGCOS.

Additionally, we did some pre-processing specifically prior to the Andersen LR test if there were inappropriate response patterns. The responses are divided into two sets based on the demographic variable. If a response option is never endorsed, then that gap is filled by left shifting all higher response options. Similarly, if one of the two sets lacks responses for the highest response option, then the other set has their highest two response options merged. This preprocessing is only done when computing the Andersen LR test.

An additional note on the collapse of response categories is for the GOS subscale (Table S 4). To mimic the original design, we turned GOS into a 5-point Likert scale. Response options 2 and 3 were merged as well as response options 5 and 6. In comparison, the subscales we designed are by default 4-point Likert scales.

The figures, tables, and values in the main paper are results obtained without imputation. We also calculated the same with imputation (not shown) as a possible way to make use of more data, but we observed minimal differences.

**Supplementary Tables /Figures**

***Table S 1. Item content for the Swedish Autism mGCOS-24.***

| Item | Subscale | Swedish text | English translation |
|---|---|---|---|
| 1 | 1 Knowledge | Jag förstår varför jag/min familj besöker den genetiska mottagningen | I understand why I/my family am/are attending the clinical genetics service |
| 2 | 4 Cognitive control | Jag kan förklara för personer i min familj vad neuropsykiatriska funktionedsättning innebär om de behöver veta. | I can explain the meaning of my neurodevelopmental condition to people in my family if they need to know. |
| 3 | 4 Cognitive control | Jag är medveten om vilken påverkan tillståndet kan ha på mitt/mina (eventuellt framtida) barn | I understand the impact of the condition may have on my (possible future) child/children |
| 4 | No subscale | Jag blir bekymrad när jag tänker på tillståndet jag har /i min familj | I get upset when thinking about the condition that I have /in my family |
| 5 | 1 Knowledge | Jag vet vart jag ska vända mig för att få det medicinska eller psykologiska stöd som jag och/eller min familj behöver (tex förbyggande åtgärder, mediciner, behandling, kontroller) | I know where to turn to receive the medical or psychological support that I and/or my family need (e.g., preemptive efforts, medications, treatment, controls) |
| 6 | 2 Hope | Jag tycker att tillståndet jag har/i min familj har lett till något positivt | I think having the condition that I have/in my family has led to something positive |
| 7 | 2 Hope | Jag upplever att jag har kontroll över hur tillståndet påverkar min familj | I feel in control over how the condition affects my family |
| 8 | 2 Hope | Jag känner mig positiv inför framtiden | I feel positive about the future |
| 9 | 2 Hope | Jag känner att jag kan hantera att ha det tillståndet jag har / i min familj | I feel able to cope with having the condition of mine /in my family |
| 10 | 1 Knowledge | Jag vet vad jag har för nytta av de alternativ som finns tillgängliga för mig (tex genetisk testning) | I know what could be gained from the options available to me (e.g., genetic testing) |
| 11 | 3 Coping | Att ha det tillståndet (i min familj) gör mig orolig | Having this condition (in my family) makes me worried |
| 12 | 4 Cognitive control | Jag vet hur det tillståndet kan påverka mina övriga släktingar på något sätt (tex syskon, farbröder/morbröder, fastrar/mostrar, kusiner) | I know how the condition could affect my other relatives in some way (e.g., siblings, uncles, aunts, cousins) |
| 13 | 4 Cognitive control | Mina beslut rörande det tillståndet kan påverka framtiden för mitt/mina (eventuellt framtida) barn | My decisions in relation to the condition can change the future for my (possibly future) child/children |
| 14 | 4 Cognitive control | Jag förstår varför jag har en remiss till genetiska mottagningen | I understand why I have been referred to the clinical genetics service |
| 15 | 4 Cognitive control | Jag vet hur jag kan få övrigt stöd jag och/eller min familj kan behöva (t.ex. från | I know how to get other kinds of support that I and/or my family might need (e.g., from counselor, |

| | | | |
|---|---|---|---|
| | | kurator, Försäkringskassan, kommunen, ekonomiskt stöd, socialt stöd) | the Social Insurance Agency, the municipality, financial support, social support) |
| 16 | 1 Knowledge | Jag kan förklara vad det neuropsykiatriska tillståndet innebär för personer utanför familjen som kan behöva veta (t ex vänner, skola, socialtjänst, arbetsplats, habilitering) | I can explain the meaning of my neurodevelopmental condition to people outside the family that might need to know (e.g., friends, school, social services, work, habilitation) |
| 17 | 4 Cognitive control | Jag vet vad jag kan göra för att förändra hur mitt neuropsykiatriska tillstånd påverkar mig eller mitt/mina (eventuellt framtida) barn | I know what I can do to change how my neurodevelopmental condition affects me or my (possibly future) child/children |
| 18 | 4 Cognitive control | Jag vet vilka i familjen som har ökad chans för att utveckla neuropsykiatriska tillståndet | I know who in the family are more likely to develop the neurodevelopmental condition |
| 19 | No subscale | Jag tror att mitt/mina (eventuellt framtida) barn kan få ett så gott liv som möjligt | I believe my (possibly future) child/children can live as rewarding life as possible |
| 20 | 2 Hope | Jag kan planera för framtiden | I am able to make plans for the future |
| 21 | 3 Coping | Jag har dåligt samvete för att jag eventuellt kan föra neuropsykiatriska tillståndet vidare till mitt/mina (eventuellt framtida) barn | I feel bad that I might pass my neurodevelopmental condition on to my (possibly future) child/children |
| 22 | 3 Coping | Jag känner mig maktlös att påverka något angående det tillståndet i min familj | I feel powerless to do anything about the condition in my family |
| 23 | No subscale | Jag förstår varför jag har kontakt med genetiska mottagningen | I understand why I am in contact with the clinical genetics service |
| 24 | 1 Knowledge | Med kunskaper om mitt neuropsykiatriska tillstånd kan jag fatta beslut som kan påverka framtiden för mitt/mina (eventuellt framtida) barn | With knowledge about my neurodevelopmental condition, I can make decisions that can change the future for my (possibly future) child/children |

***Table S 2 Item-level Rasch fit statistics for the Swedish Autism mGCOS-24.*** *MSQ calculations are based on conditional calculations (n = 155 complete cases). Simulation based thresholds are from 71 simulated datasets.*

| Item | InfitMSQ | Infit thresholds | Infit diff |
|---|---|---|---|
| Q1 | 1.119 | [0.896, 1.098] | 0.021 |
| Q2 | 0.963 | [0.861, 1.154] | no misfit |
| Q3 | 0.949 | [0.853, 1.168] | no misfit |
| Q4 | 1.264 | [0.866, 1.105] | 0.159 |
| Q5 | 1.04 | [0.915, 1.098] | no misfit |
| Q6 | 0.917 | [0.929, 1.175] | 0.012 |
| Q7 | 0.827 | [0.868, 1.133] | 0.041 |
| Q8 | 0.866 | [0.912, 1.192] | 0.046 |
| Q9 | 0.813 | [0.948, 1.169] | 0.135 |
| Q10 | 0.842 | [0.852, 1.123] | 0.01 |
| Q11 | 1.264 | [0.811, 1.208] | 0.056 |
| Q12 | 1.06 | [0.913, 1.138] | no misfit |
| Q13 | 1.316 | [0.866, 1.092] | 0.224 |
| Q14 | 0.968 | [0.82, 1.106] | no misfit |
| Q15 | 0.779 | [0.854, 1.093] | 0.075 |
| Q16 | 0.801 | [0.839, 1.124] | 0.038 |
| Q17 | 0.8 | [0.861, 1.13] | 0.061 |
| Q18 | 1.15 | [0.873, 1.127] | 0.023 |
| Q19 | 0.96 | [0.885, 1.2] | no misfit |
| Q20 | 0.937 | [0.757, 1.054] | no misfit |
| Q21 | 1.361 | [0.908, 1.118] | 0.243 |
| Q22 | 1.106 | [0.854, 1.129] | no misfit |
| Q23 | 0.983 | [0.884, 1.083] | no misfit |
| Q24 | 0.958 | [0.899, 1.17] | no misfit |

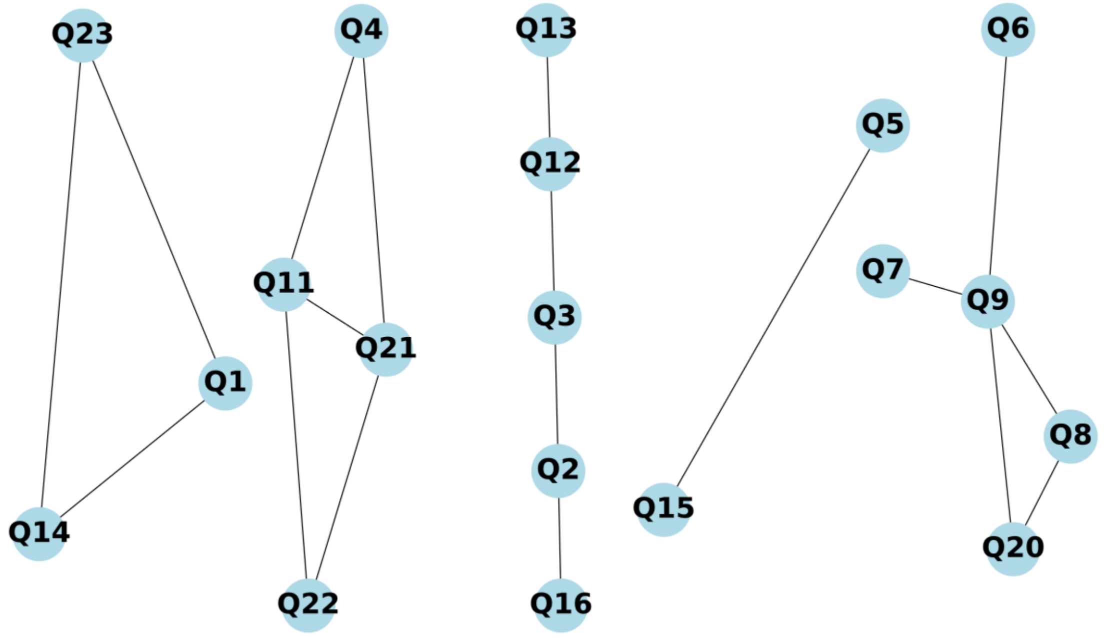


***Figure S 1. Residual correlation network for the Swedish Autism mGCOS-24.*** *Graphical visualization of local item dependencies, where edges indicate residual correlations exceeding simulation-based cutoffs.*

***Table S 3. Item-level Rasch fit statistics for the Swedish General GCOS-24.*** *MSQ calculations are based on conditional calculations (n = 344 complete cases). Simulation based thresholds are from 58 simulated datasets.*

| Item | InfitMSQ | Infit thresholds | Infit diff |
|---|---|---|---|
| Q1 | 1.015 | [0.924, 1.071] | no misfit |
| Q2 | 0.898 | [0.913, 1.074] | 0.015 |
| Q3 | 0.909 | [0.896, 1.107] | no misfit |
| Q4 | 1.239 | [0.86, 1.052] | 0.187 |
| Q5 | 0.973 | [0.887, 1.109] | no misfit |
| Q6 | 1.242 | [0.888, 1.079] | 0.163 |
| Q7 | 0.837 | [0.966, 1.086] | 0.129 |
| Q8 | 0.832 | [0.877, 1.058] | 0.045 |
| Q9 | 0.759 | [0.931, 1.097] | 0.172 |
| Q10 | 0.791 | [0.89, 1.208] | 0.099 |
| Q11 | 1.202 | [0.899, 1.114] | 0.088 |
| Q12 | 0.973 | [0.848, 1.06] | no misfit |
| Q13 | 1.264 | [0.909, 1.133] | 0.131 |
| Q14 | 1.052 | [0.906, 1.092] | no misfit |
| Q15 | 0.977 | [0.898, 1.028] | no misfit |
| Q16 | 0.792 | [0.883, 1.109] | 0.091 |
| Q17 | 0.841 | [0.869, 1.079] | 0.028 |
| Q18 | 1.121 | [0.875, 1.157] | no misfit |
| Q19 | 0.892 | [0.919, 1.14] | 0.027 |
| Q20 | 0.887 | [0.902, 1.109] | 0.015 |
| Q21 | 1.451 | [0.915, 1.123] | 0.328 |
| Q22 | 1.192 | [0.949, 1.132] | 0.06 |
| Q23 | 1.013 | [0.943, 1.15] | no misfit |
| Q24 | 1.135 | [0.883, 1.047] | 0.088 |

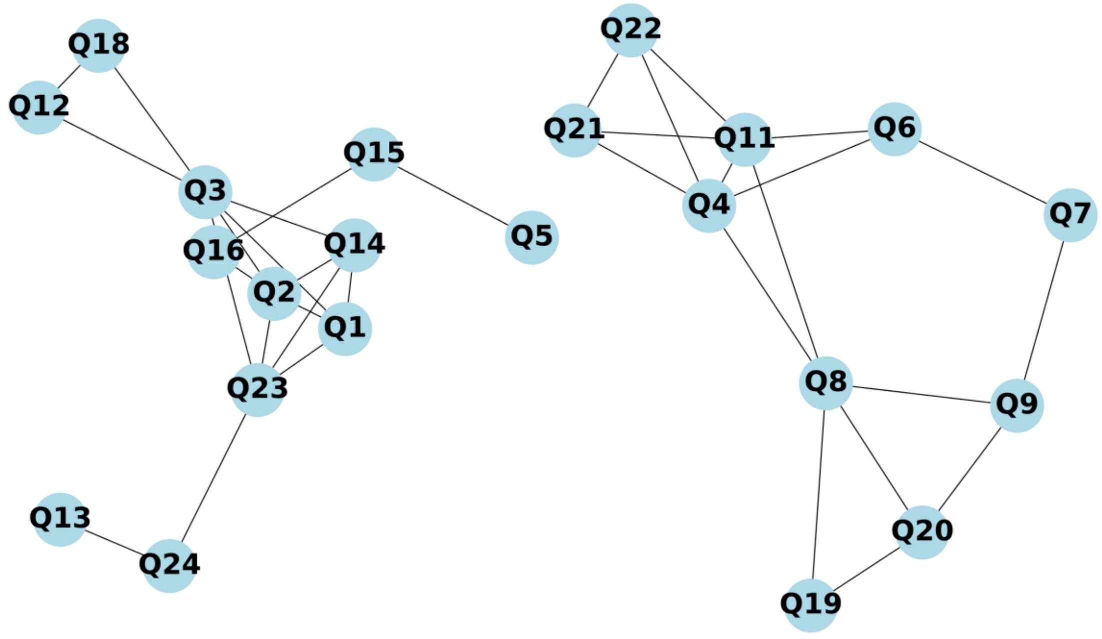


***Figure S 2. Residual correlation network for the Swedish General GCOS-24.*** *An edge between two items represents an above-cutoff correlation between the two items.*

***Table S 4. Pairwise subscale combinations and Genomic Outcome Scale (GOS) evaluation***
*Results showing that combining subscales or evaluating the GOS yields multidimensionality.*

| **Subscales (autism mGCOS)** | **1st Eigenvalue** | **Eigenvalue Threshold** |
|---|---|---|
| GOS | 1.87 | 1.63 |
| Subscale 1 + Subscale 2 | 2.18 | 1.78 |
| Subscale 1 + Subscale 3 | 2.86 | 1.72 |
| Subscale 1 + Subscale 4 | 1.89 | 1.78 |
| Subscale 2 + Subscale 3 | 2.54 | 1.62 |
| Subscale 2 + Subscale 4 | 3.05 | 1.74 |
| Subscale 3 + Subscale 4 | 3.30 | 1.81 |
| **Subscales (general GCOS)** | **1st Eigenvalue** | **Eigenvalue Threshold** |
| GOS | 1.61 | 1.48 |

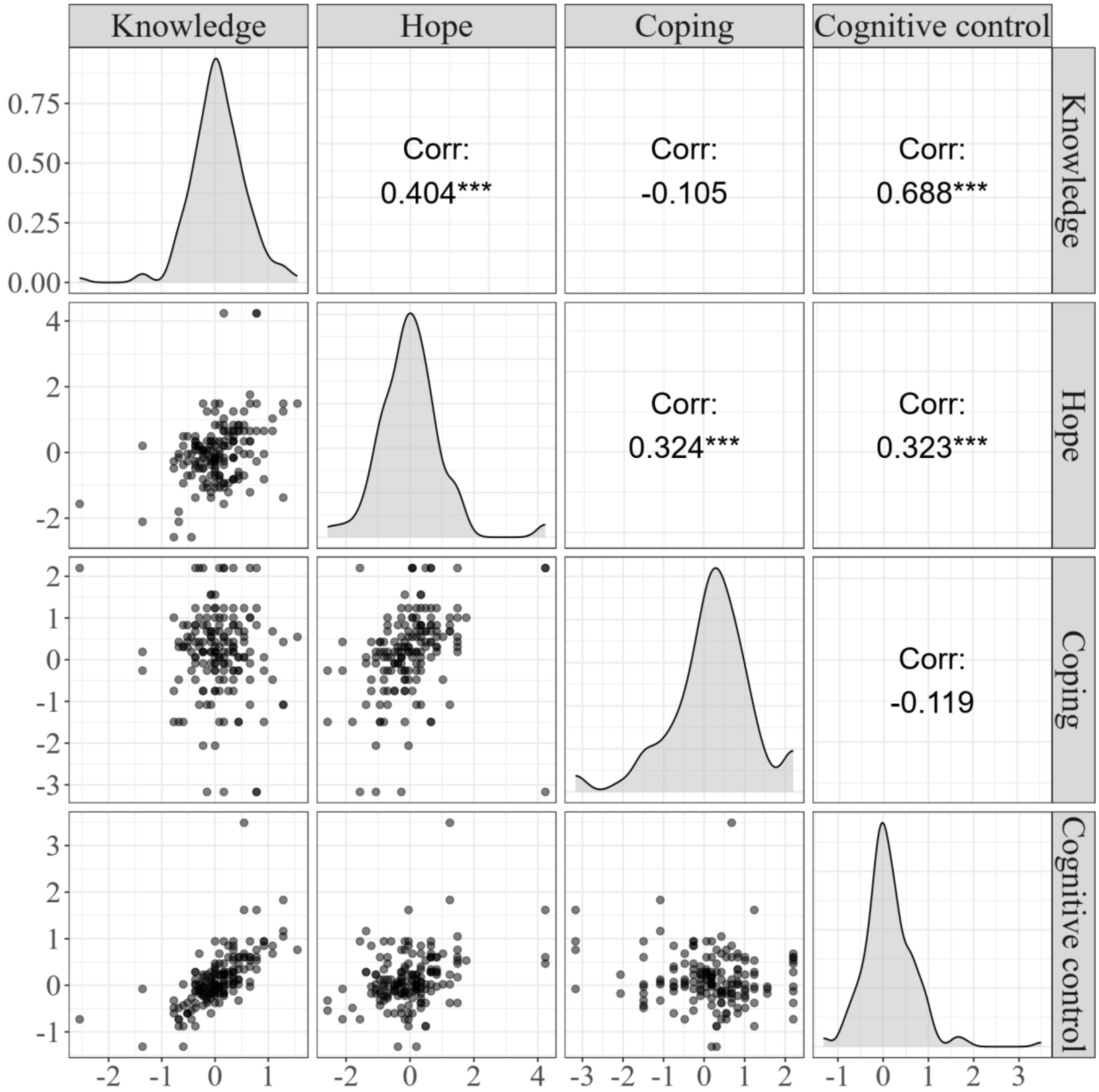


***Figure S 3, person locations and their correlations between subscales for the Swedish Autism mGCOS.*** *Correlations range from no significance to medium-high Pearson's r. Scatter plots show individuals subscale-pairs of person location. Participants included were the 161 who responded to at least 2 items on each subscale. *** p < 0.001.*

***Table S 5. Fit statistics for the GOS applied to the Swedish General GCOS-24.*** *MSQ calculations are based on conditional calculations (n = 331 complete cases). Simulation based thresholds are from 100 simulated datasets.*

| Item | InfitMSQ | Infit thresholds | Infit diff |
|---|---|---|---|
| Q16 | 0.889 | [0.93, 1.134] | 0.041 |
| Q18 | 1.016 | [0.895, 1.112] | no misfit |
| Q4 | 1.319 | [0.872, 1.085] | 0.234 |
| Q17 | 0.781 | [0.898, 1.124] | 0.117 |
| Q20 | 0.994 | [0.891, 1.135] | no misfit |
| Q24 | 1.135 | [0.899, 1.165] | no misfit |
